# Wearable alcohol monitors for alcohol use data collection among college students: feasibility and acceptability in a pilot study

**DOI:** 10.1101/2021.02.17.21251959

**Authors:** Molly Rosenberg, Christina Ludema, Sina Kianersi, Maya Luetke, Kristen Jozkowski, Lucia Guerra-Reyes, Patrick C. Shih, Peter Finn

## Abstract

**Objective:** To assess the feasibility and acceptability of using BACtrack Skyn wearable alcohol monitors in a college student population.

**Method:** In September 2019, we enrolled n=5 Indiana University undergraduate students in a study to wear alcohol monitor wristbands continuously over a 5-day period. Concurrently, participants completed daily surveys querying details about their alcohol use in the previous 24 hours. We measured acceptability at endline with the Acceptability of Intervention Measure (AIM) scale (min=1, max=5). We measured feasibility with process measures: 1) amount of alcohol monitor data produced, and 2) correlation between drinking events identified by the alcohol monitors and drinking events reported by participants.

**Result:** Participants reported high acceptability of the wearable alcohol monitors with a mean AIM score of 4.3 (range: 3.3 to 5.0). Feasibility of monitor use was high: A total of 589 hours of alcohol use data was collected. All participants were able to successfully use the alcohol monitors, producing a total of 24 out of 25 possible days of alcohol monitoring data. Participants reported a total of 15 drinking events during follow-up and we detected 12 drinking events with the alcohol monitors. The self-reported drinking event start times were highly correlated with the monitor detected event start time (Spearman’s ρ=0.9, p<0.0001). The self-reported number of drinks during a drinking event was correlated with the area under the curve of each drinking event peak (Pearson’s r=0.7, p=0.008).

**Conclusion:** Wearable alcohol monitors are a promising data collection tool for more objective real-time measures of alcohol use in college student populations.

## Introduction

College students are at particularly high risk for heavy alcohol consumption and alcohol-related harms. Overall, approximately 75% of college students report alcohol use in the last year and 60% within the last month (Schulenberg et al., 2019). Further, as many as 28% of college students report binge drinking (i.e. consuming five or more drinks in a row) in the previous 2 weeks (Schulenberg, et al., 2019). Heavy alcohol consumption among college students has been consistently linked to adverse outcomes such as academic difficulties, injuries, and death (Fairlie et al., 2019; Hingson et al., 2017; Patrick et al., 2016). Alcohol consumption is also associated with numerous sexual risks in this population, including sexual assault victimization and perpetration (Abbey, 2011; Brown & Vanable, 2007; Cooper, 2002; Crane et al., 2016; Davis et al., 2014; Mohler-Kuo et al., 2004). Despite the clear risks associated with alcohol use among college students, our understanding of the epidemiology of alcohol use and alcohol-related harms on college campuses is limited.

Indeed, researchers have historically been reliant on self-reports to measure alcohol use among college students. Self-reported alcohol use data are limited by the potential for social desirability bias and recall bias, the latter of which is heightened in cases of alcohol-induced memory loss (Rose & Grant, 2010). Objective and passively collected data on alcohol use in college students would improve our understanding of the campus alcohol landscape and improve our ability to identify effective alcohol risk reduction interventions.

There has thus been sustained interest in developing novel and objective ways to passively collect alcohol use data. Recent decades have seen the development and evaluation of several wearable monitors that measure transdermal alcohol concentration (TAC) (Swift, 2000), including the Secure Continuous Remote Alcohol Monitor (SCRAM) (Karns-Wright et al., 2018) and the Wrist Transdermal Alcohol Sensor (WrisTAS) (Sirlanci et al., 2019). Both devices have shown good correlation between self-reported alcohol consumption and TAC data from the sensors (Alessi et al., 2019; Fairbairn et al., 2019; Karns-Wright, et al., 2018; Rash et al., 2019; Simons et al., 2015). However, both the SCRAM (worn on the ankle) and the WrisTAS (worn on the wrist) are fairly obtrusive, noticeable devices, and have largely been used to monitor alcohol abstinence (Karns-Wright, et al., 2018; Luczak & Ramchandani, 2019). In particular, the SCRAM, which is commonly used in the criminal justice system, resembles an ankle GPS tracking device worn by people who are incarcerated, a similarity which may influence willingness to wear such a device in a field-based research context.

The next generation of wearable alcohol monitors includes a device (‘Skyn’) developed by BACTrack©. The BACTrack Skyn has a TAC sensor that is integrated into a small, unobtrusive wristband, and may address some of the limitations of prior models (Campbell et al., 2018; Fairbairn & Kang, 2019; Wang et al., 2019). Because this technology is newly developed, it is not yet known whether college students are able and willing to wear the devices for research purposes, and whether the devices would provide data consistent with real-life college drinking events. As such, the aim of this pilot study was to assess the feasibility and acceptability of using BACTrack Skyn wearable alcohol monitors in a college student population.

## Method

### Study population

To test the feasibility and acceptability of the novel wearable alcohol monitors in a college student population, we enrolled n=5 undergraduate Indiana University students to wear the devices continuously over a five day study period. Participants were eligible to enroll in the study if they were aged 21 years or older, English-speaking, and consuming 1 or more alcoholic drinks per week at the time of the study. Participants were recruited using on-campus and on-line flyers to publicize the study. Interested participants contacted study staff who confirmed eligibility and scheduled a baseline visit to obtain written informed consent. Participants were paid $55 for completion of all study procedures. Ethical approval of the study protocol was provided by the Indiana University Human Subjects Office (#1907111038).

### Study procedures

All participants were asked to wear a ‘BACtrack Skyn’ alcohol monitor for five days. The BACtrack Skyn monitors are wristbands equipped with a small sensor to measure transdermal alcohol content (TAC) with measurements recorded approximately every 20 seconds. The sensor pairs via Bluetooth to a smartphone application downloaded onto the participants phone at the baseline study visit. The TAC data are then transmitted from the mobile app to a secure server maintained by BACtrack. During the baseline visit, study staff trained participants to properly use the monitors and instructed them to wear the devices continuously for five days, except for removal for regular charging (at least every other day), and when showering or swimming, because the devices are not waterproof. Participants were asked to complete a baseline survey to provide socio-demographic and behavioral data, short daily surveys querying the drinking and sexual behaviors of the previous day, and an endline survey focused on participant acceptability of the study procedures. The baseline and endline surveys were completed during each of the two study visits. Links to the daily surveys were pushed to participants’ mobile phones every day at 1pm and participants completed them remotely. We did not make the alcohol use data available to the study participants during follow-up to minimize the potential for negative reactivity, but provided them with a summary of their alcohol use data at the endline visit.

### Key measures

We measured acceptability with the 4-item *Acceptability of Intervention Measure (AIM) scale*, collected in the endline survey.(Weiner et al., 2017) We queried the AIM separately for the alcohol monitor use, the daily surveys, and regarding a hypothetical future intervention using data from alcohol monitors to inform sexual decision-making. We measured feasibility with the *Feasibility of Intervention Measure (FIM) scale*, collected in the endline survey (Weiner et al., 2017). We queried the 4-item FIM separately for the alcohol monitor use and the daily surveys. Each of the four AIM and FIM item responses are measured on a five-point Likert scale. The scores are calculated as the mean of the four item responses for a range from 1 to 5.

We further measured feasibility with the following process measures:

1. *The amount of alcohol data produced* by the study participants wearing the alcohol monitors, measured by the number of TAC data points collected, and number of days with recorded TAC data (out of a total of 25 possible days).
2. *The number of daily surveys completed* by the participants, out of a total of 25 possible surveys.
3. *The correlation between the drinking event start time self-reported by the participants and identified by monitor*. In the daily surveys, participants reported the number of standard drinks they consumed in the previous 24 hours. Participants who reported >0 drinks were considered to have had a self-reported drinking event, and were queried about the time of day the drinking event began. We processed the TAC data to identify drinking events and derive the associated event start times. Processing the data is necessary because the TAC data are in the form of a signal with noise with no natural zero starting value, and baseline TAC varies between individuals (Fairbairn & Kang, 2019). We used median filtering and moving average filtering to remove the noise from the data. After processing, we identified peaks (i.e. drinking events) in the processed TAC signal using two of the peak properties, peak width and peak prominence. We set value ranges for these properties to maximize the peak detection sensitivity and specificity, setting the width range to 117 to 289 minutes (350 to 866 data points), and the minimum prominence value to 3.4 TAC units. A sensitivity analysis assessing differences in drinking event detection for lower minimum prominence values was conducted and results presented in supplemental Table A.1. We identified the start and end time of a drinking event as the left bases and right bases of the peaks, respectively. We tested the strength of the correlation between the start times of self-reported and monitor-identified drinking events by calculating the Spearman correlation coefficient. The correlation coefficient was determined to be significantly different from zero if its associated p-value was <0.05.
4. *The correlation between the number of standard drinks self-reported in each drinking event and the area under the curve (AUC) of each TAC drinking event peak from the alcohol monitors*. Participants self-reported the number of standard drinks consumed in the previous 24 hours in the daily survey. Available responses were: 0, 1-2, 3-4, 5-6, and 7+. For analysis, we recoded these responses at the midpoint of the response category (e.g. category ‘1-2’ was recoded to 1.5 drinks), and recoded the ‘7+’ response to 7. We calculated the AUC for each TAC peak using the estimated drinking event start time and end time as described above. The AUC was calculated as the area between the start and end time, i.e. left and right bases of a peak (Figure 1). We tested the strength of the correlation between the number of self-reported drinks per drinking event and the monitor-produced area under the TAC curve by calculating the Pearson correlation coefficient. The correlation coefficient was determined to be significantly different from zero if its associated p-value was <0.05. **Figure 1.**
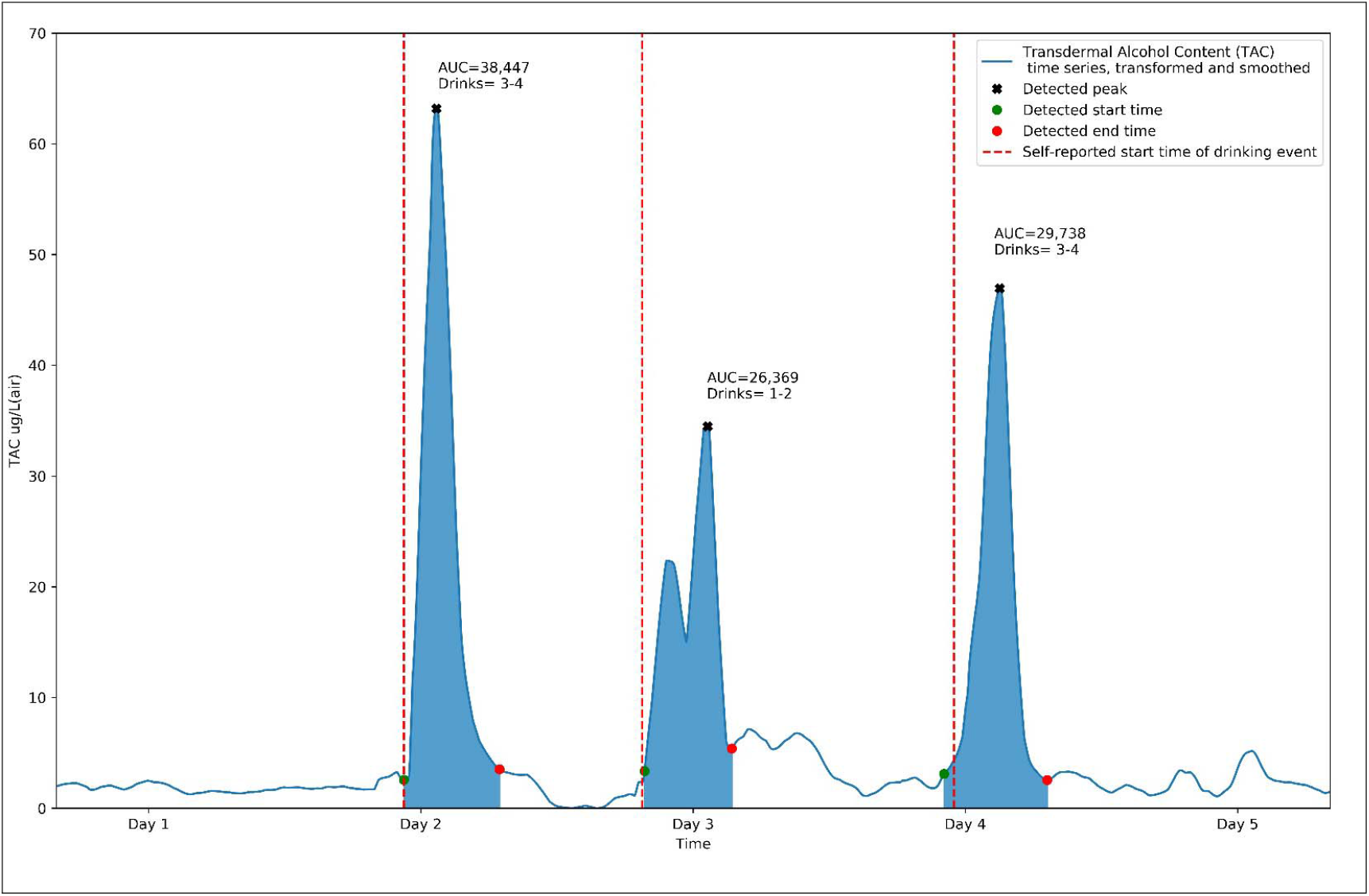
Example alcohol monitor data produced by pilot participant over 5 days, annotated with self-reported drinking events and drinking amounts, September 2019. For this correlation analysis, we treated each drinking event as an independent event. To assess whether this assumption was valid given that multiple drinking events were reported by each participant, we conducted a sensitivity analysis calculating the within-person correlation between number of drinks and AUC for participants with at least three drinking events (n=2). The correlation coefficients for these two participants were similar in direction and magnitude to the overall correlation coefficient across all drinking events (data not shown).

We used the SciPy library in Python for both peak detection and correlation calculations (see supplemental Table A.2 for summary of packages used) (Jones et al., 2001).

## Results

Overall, we enrolled five participants in this pilot study (Table 1). Two were male-identified and three were female-identified. The mean age was 21.6 years and ranged from 21.2 to 22.3 years. The sample was diverse with respect to racial/ethnic identity, with White, Hispanic, Black/African-American, and Asian participants represented. At baseline, all participants reported an average alcohol consumption frequency of 3-5 times per week, with most reporting an average number of standard drinks per drinking event between 3 to 4 drinks.

**Table 1.** Pilot study participant socio-demographic and behavioral characteristics at baseline, n=5, September 2019

| Characteristic | N (%) |
| --- | --- |
| Gender* |  |
| Male | 2 (40%) |
| Female | 3 (60%) |
| Mean age (range) | 21.6 (21.2 – 22.3) |
| Racial identity |  |
| White | 2 (40%) |
| Hispanic/Latinx | 1 (20%) |
| Black/African American | 1 (20%) |
| Asian | 1 (20%) |
| School year |  |
| 3<sup>rd</sup> year undergraduate | 2 (40%) |
| 4<sup>th</sup> year undergraduate | 2 (40%) |
| 5<sup>th</sup>+ year undergraduate | 1 (20%) |
| Greek affiliated |  |
| Yes | 1 (20%) |
| No | 4 (80%) |
| Alcohol consumption frequency |  |
| Daily | 0 (0%) |
| 3-5 times a week | 5 (100%) |
| Once a week or less | 0 (0%) |
| Average # drinks per event |  |
| 1-2 | 1 (20%) |
| 3-4 | 3 (60%) |
| 5-6 | 1 (20%) |

\*All participants reported concordance between their sex at birth and current gender identity

Participants reported high levels of acceptability with and feasibility regarding the study procedures in the endline survey (Table 2). The average AIM scale scores were 4.3 and 4.4 for the alcohol monitor use and daily surveys, respectively. Participants also reported high acceptability for a future intervention using data from the alcohol monitors to inform sexual decision-making (mean AIM 4.5). The average FIM scale scores were in a similarly high range for the alcohol monitor use (4.3) and daily surveys (4.7).

**Table 2.** Acceptability and feasibility scale data, n=5 pilot participants after 5 days of follow-up

| Scale | Component | Mean | Range |
| --- | --- | --- | --- |
| Acceptability of Intervention Measure | Alcohol monitor use | 4.3 | 3.3 – 5.0 |
|  | Daily surveys | 4.4 | 3.8 – 5.0 |
|  | Future intervention | 4.5 | 3.5 – 5.0 |
| Feasibility of Intervention Measure | Alcohol monitor use | 4.3 | 3.3 – 5.0 |
|  | Daily surveys | 4.7 | 4.3 – 5.0 |

Feasibility was further supported by the amount of data collected over the study period (Table 3). We collected a total of 106,099 TAC datapoints, covering an equivalent of 589 hours. Of the total 25 possible days of alcohol monitor data (i.e. five days for each of the five participants), we collected data in 24 days. The single day in which data were not collected was due to a device’s battery running out of charge, not because of noncompliance on the participant’s part. Each participant returned all five daily surveys for a total of 25 daily surveys (100% returned). Participant-level data on feasibility, acceptability, and use experiences are reported in supplemental Table A.3.

**Table 3.** Feasibility process measures, n=5 participants over 5 days of follow-up

| **Feasibility item** | **Quantitative measure** |
| --- | --- |
| Overall number of collected TAC data points | 106,099 (589 hours) |
| Proportion of days of alcohol monitor data produced | 24/25 (96%) |
| Proportion of daily surveys returned | 25/25 (100%) |
| Number of drinking events, self-reported | 15 |
| Number of drinking events, monitor identified | 12 |
| Correlation between drinking event start time self-reported and identified by the monitors (p-value) | 0.90 (<.0001) |
| Correlation between estimated # drinks and area under the TAC curve (p-value) | 0.72 (0.008) |

The alcohol monitors produced data that were correlated with the drinking event behavior self-reported by the participants (Table 3). Participants self-reported a total of 15 drinking events and we identified 12 drinking events from the alcohol monitor data. One false positive drinking event was detected in the TAC data, occurring the same day as a correctly detected self-reported event. Characteristics of drinking events detected and not detected by the monitors are presented in supplemental Table A.4. The Spearman correlation between the self-reported and alcohol monitor derived start time of the drinking event was 0.90 (p-value < .0001). The Spearman correlation between the self-reported number of drinks per drinking event and the area under the TAC curve for the drinking event was 0.7 (p-value: 0.008). Figure 1 provides example data from a pilot participant over the five days of follow-up. Visual inspection of the figure further underscores the correlation between the drinking event start-time from the TAC and self-reported data, and the correlation between the peak size and number of drinks self-reported.

## Discussion

Overall, we found high acceptability and feasibility of using BACTrack Skyn wearable alcohol monitors to measure drinking behaviors among college students. Participants in this pilot study were able to use the alcohol monitors to produce nearly complete continuous TAC data over five days of follow-up, and characteristics derived from the TAC dataset were largely reflective of the self-reported timing and magnitude of drinking events. Participants also reported high compliance with and acceptability of the daily survey procedures necessary to collect the event-level self-reported data.

The BACTrack Skyn alcohol monitor is a newly developed biosensor, so few studies have assessed its use and acceptability for continuous transdermal alcohol monitoring. To our knowledge, the two published validity studies used a prototype version of the device and were conducted in laboratory settings only (Fairbairn & Kang, 2019; Wang, et al., 2019). In qualitative assessments, one of these studies found that the devices were generally acceptable to participants (Wang, et al., 2019). Our study is the first to demonstrate acceptability and preliminary validity in the field and among college students. The BACTrack Skyn device has been developed to be sleeker than its predecessors, and more closely resemble existing fitness trackers on the market (Fairbairn & Kang, 2019; Wang, et al., 2019). The high levels of acceptability reported by our college student participants is consistent with the fact that young US adults aged 18 to 34 years have the highest levels of engagement with wearable fitness devices of all age groups, with nearly 30% of this age group reporting current use of a wearable fitness tracker (McCarthy, 2019).

The TAC data processing methods we used detected many, but not all, drinking events self-reported by participants. We were only unable to identify a total of three drinking events of the 15 reported by participants. All three of these events were reported by a single participant and comprised events during which low levels of alcohol consumption were reported (1-2 drinks). It is likely that this particular participant had technological challenges (e.g. infrequent charging) that the other participants did not experience. It is also possible that low levels of alcohol consumption for this particular participant did not produce large enough TAC peaks to be detected as a drinking event with our peak detection method. Future studies should incorporate more structured endline interviews about user experiences (e.g. charging, position of band), and user characteristics (e.g. body mass index, perspiration levels, drinking event magnitude) to assess more definitively their potential influence on data accuracy. Peak detection methods can be modified to maximize detection of alcohol consumption events (at the expense of increasing false positives) or minimize detection of events that are not substantiated by self-report (at the expense of missing low-peak events), as shown in our sensitivity analysis. Calibration of these peak identification methods will likely be specific to the goal of a particular research project and are an area for future research (Kianersi et al., 2020; Roache et al., 2019).

Notably, only one drinking event was detected in our TAC data that was not self-reported by the participants. However, it is important to note that the standard to which we compared the TAC data was participant self-report, with the potential for recall error and social desirability bias. A specific limitation of our self-reported data was the lack of precision our binned response categories provided for the number of self-reported drinks consumed (e.g. ‘1-2’ and ‘3-4’). More exact numbers would have provided a stronger comparison and should be elicited in future surveys. A further limitation arises from the time lag between the drinking event and the self-reported time of drinking initiation. Although the recall period is relatively short – participants reported on the previous day’s drinking behaviors the next day – the accuracy of self-reported data would likely improve with even shorter time lags or real-time data collection. Though beyond the scope of this current study, future validity studies should consider incorporating a laboratory component or use breathalyzer data as more rigorous ‘gold standard’ comparisons.

Our small sample size and convenience sampling technique indicate that our results should be interpreted with caution. We enrolled a very small number of participants (n=5) who self-selected into this pilot study with five days of follow-up. Thus, the statistical inferences we can make are limited and our findings are unlikely to be representative of all college students or the diversity of their drinking experiences over time. That said, because of the continuous data collection from the alcohol monitors, the five days of follow-up produced a large amount of TAC data (nearly 600 hours) which we were able to analyze at the drinking event level (n=15) as opposed to the individual level (n=5). Nonetheless, future studies should enroll a larger college student study population using a random sampling technique to confirm our feasibility and acceptability findings.

### Conclusions and future directions

Our findings underscore the promise of using BACTrack Skyn wearable alcohol monitor technology to improve our understanding of alcohol consumption among college students, a population at particularly high risk for alcohol-related harms. Future applications of this new technology have the potential to overcome some of the biases that arise from traditional self-reported alcohol consumption data and could be leveraged to identify more precise recommendations on the levels of alcohol consumption that confer increased risk for specific alcohol-related harms. Given the high levels of acceptability demonstrated in our study, future studies could also consider using these alcohol monitors as a platform to deliver interventions based on real-time alcohol consumption or to give personalized alcohol consumption feedback to users.

## Data Availability

Data and code are available upon request.

## Acknowledgements

The authors are exceedingly grateful to all those involved in the data collection procedure, including Finn Lab staff Lindsay Fisher and Eli Farmer, and, most importantly, the study participants themselves. The authors declare no conflict of interest.

## Data availability

The data underlying this article will be shared on reasonable request to the corresponding author.

## Notes

**Role of the funding source:** The funding sources had no involvement in study design; in the collection, analysis and interpretation of data; in the writing of the report; nor in the decision to submit the article for publication.

**Funding:** This work was supported by the Vice Provost for Research through the Research Equipment Fund; the NIH/NIAAA grant [R01 AA13650 to Peter Finn]

### Competing Interest Statement

The authors have declared no competing interest.

### Funding Statement

This work was supported by the Vice Provost for Research through the Research Equipment Fund; the NIH/NIAAA grant [R01 AA13650 to Peter Finn]

### Author Declarations

Ethical approval of the study protocol was provided by the Indiana University's Human Subjects & Institutional Review Boards (Protocol #1907111038).

## REFERENCES

Abbey, A. (2011). Alcohol’s role in sexual violence perpetration: Theoretical explanations, existing evidence and future directions. Drug and alcohol review, 30(5), 481–489.

Alessi, S. M., Barnett, N. P., & Petry, N. M. (2019). Objective continuous monitoring of alcohol consumption for three months among alcohol use disorder treatment outpatients. Alcohol, 81, 131–138.

Brown, J. L., & Vanable, P. A. (2007). Alcohol use, partner type, and risky sexual behavior among college students: Findings from an event-level study. Addictive Behaviors, 32(12), 2940–2952. doi:10.1016/j.addbeh.2007.06.011

Campbell, A. S., Kim, J., & Wang, J. (2018). Wearable electrochemical alcohol biosensors. Current opinion in electrochemistry, 10, 126–135.

Cooper, M. L. (2002). Alcohol use and risky sexual behavior among college students and youth: Evaluating the evidence. Journal of Studies on Alcohol, 101–117. doi:DOI 10.15288/jsas.2002.s14.101

Crane, C. A., Godleski, S. A., Przybyla, S. M., Schlauch, R. C., & Testa, M. (2016). The proximal effects of acute alcohol consumption on male-to-female aggression: a meta-analytic review of the experimental literature. Trauma, Violence, & Abuse, 17(5), 520–531.

Davis, K. C., Masters, N. T., Eakins, D., Danube, C. L., George, W. H., Norris, J., & Heiman, J. R. (2014). Alcohol intoxication and condom use self-efficacy effects on women’s condom use intentions. Addictive Behaviors, 39(1), 153–158. doi:10.1016/j.addbeh.2013.09.019

Fairbairn, C. E., & Kang, D. (2019). Temporal Dynamics of Transdermal Alcohol Concentration Measured via New-Generation Wrist-Worn Biosensor. Alcoholism: Clinical and Experimental Research, 43(10), 2060–2069.

Fairbairn, C. E., Rosen, I. G., Luczak, S. E., & Venerable, W. J. (2019). Estimating the quantity and time course of alcohol consumption from transdermal alcohol sensor data: A combined laboratory-ambulatory study. Alcohol, 81, 111–116.

Fairlie, A. M., Cadigan, J. M., Patrick, M. E., Larimer, M. E., & Lee, C. M. (2019). Unplanned Heavy Episodic and High-Intensity Drinking: Daily-Level Associations With Mood, Context, and Negative Consequences. J Stud Alcohol Drugs, 80(3), 331–339.

Hingson, R., Zha, W. X., & Smyth, D. (2017). Magnitude and Trends in Heavy Episodic Drinking, Alcohol-Impaired Driving, and Alcohol-Related Mortality and Overdose Hospitalizations Among Emerging Adults of College Ages 18-24 in the United States, 1998-2014. Journal of Studies on Alcohol and Drugs, 78(4), 540–548. doi:DOI 10.15288/jsad.2017.78.540

Jones, E., Oliphant, T., & Peterson, P. (2001). SciPy: Open source scientific tools for Python.

Karns-Wright, T. E., Dougherty, D. M., Hill-Kapturczak, N., Mathias, C. W., & Roache, J. D. (2018). The correspondence between transdermal alcohol monitoring and daily self reported alcohol consumption. Addictive Behaviors, 85, 147–152. doi:10.1016/j.addbeh.2018.06.006

Kianersi, S., Luetke, M., Agley, J., Gassman, R., Ludema, C., & Rosenberg, M. (2020). Validation of transdermal alcohol concentration data collected using wearable alcohol monitors: A systematic review and meta-analysis. Drug and Alcohol Dependence, 108304.

Luczak, S. E., & Ramchandani, V. A. (2019). Special issue on alcohol biosensors: Development, use, and state of the field: Summary, conclusions, and future directions. Alcohol (Fayetteville, NY), 81, 161.

McCarthy, J. (2019). One in Five U.S. Adults Use Health Apps, Wearable Trackers. Retrieved from https://news.gallup.com/poll/269096/one-five-adults-health-apps-wearable-trackers.aspx

Mohler-Kuo, M., Dowdall, G. W., Koss, M. P., & Wechsler, H. (2004). Correlates of rape while intoxicated in a national sample of college women. Journal of studies on alcohol, 65(1), 37–45.

Patrick, M. E., Cronce, J. M., Fairlie, A. M., Atkins, D. C., & Lee, C. M. (2016). Day-to-day variations in high-intensity drinking, expectancies, and positive and negative alcohol-related consequences. Addict Behav, 58, 110–116. doi:10.1016/j.addbeh.2016.02.025

Rash, C. J., Petry, N. M., Alessi, S. M., & Barnett, N. P. (2019). Monitoring alcohol use in heavy drinking soup kitchen attendees. Alcohol, 81, 139–147.

Roache, J. D., Karns-Wright, T. E., Goros, M., Hill-Kapturczak, N., Mathias, C. W., & Dougherty, D. M. (2019). Processing transdermal alcohol concentration (TAC) data to detect low-level drinking. Alcohol, 81, 101–110.

Rose, M. E., & Grant, J. E. (2010). Alcohol-induced blackout: phenomenology, biological basis, and gender differences. Journal of addiction medicine, 4(2), 61–73.

Schulenberg, J. E., Johnston, L. D., O’Malley, P. M., Bachman, J. G., Miech, R. A., & Patrick, M. E. (2019). Monitoring the Future national survey results on drug use, 1975–2018: Volume II, College students and adults ages 19–60. Retrieved from Ann Arbor: http://monitoringthefuture.org/pubs.html#monographs

Simons, J. S., Wills, T. A., Emery, N. N., & Marks, R. M. (2015). Quantifying alcohol consumption: Self-report, transdermal assessment, and prediction of dependence symptoms. Addict Behav, 50, 205–212. doi:10.1016/j.addbeh.2015.06.042

Sirlanci, M., Rosen, I. G., Wall, T. L., & Luczak, S. E. (2019). Applying a novel population-based model approach to estimating breath alcohol concentration (BrAC) from transdermal alcohol concentration (TAC) biosensor data. Alcohol, 81, 117–129. doi:10.1016/j.alcohol.2018.09.005

Swift, R. (2000). Transdermal alcohol measurement for estimation of blood alcohol concentration. Alcoholism, clinical and experimental research, 24(4), 422–423.

Wang, Y., Fridberg, D. J., Leeman, R. F., Cook, R. L., & Porges, E. C. (2019). Wrist-worn alcohol biosensors: strengths, limitations, and future directions. Alcohol, 81, 83–92.

Weiner, B. J., Lewis, C. C., Stanick, C., Powell, B. J., Dorsey, C. N., Clary, A. S., … Halko, H. (2017). Psychometric assessment of three newly developed implementation outcome measures. Implementation Science, 12(1), 108.

